# Hospital Variability in the Use of Vasoactive Agents in Patients Hospitalized for Acute Decompensated Heart Failure: Clinical Phenotypes and Therapeutic Approaches

**DOI:** 10.1101/2024.05.24.24307917

**Authors:** Yasuyuki Shiraishi, Nozomi Niimi, Shun Kohsaka, Kazumasa Harada, Takashi Kohno, Makoto Takei, Takahiro Jimba, Hiroki Nakano, Junya Matsuda, Akito Shindo, Daisuke Kitano, Shigeto Tsukamoto, Shinji Koba, Takeshi Yamamoto, Morimasa Takayama

**Affiliations:** Tokyo Cardiovascular Care Unit Network Scientific Committee, Tokyo, Japan; Department of Cardiology, Keio University School of Medicine, Tokyo, Japan

**Keywords:** hospital variability, heart failure, congestion, tissue hypoperfusion, vasodilator, inotrope

## Abstract

**Background:** The absence of practice standards in vasoactive agent usage for acute decompensated heart failure (ADHF) has resulted in significant treatment variability across hospitals, potentially affecting patient outcomes. This study aimed to assess temporal trends and institutional differences in vasodilator and inotrope/vasopressor utilization among ADHF patients, considering their clinical phenotypes.

**Methods:** Data were extracted from a government-funded multicenter registry covering the Tokyo metropolitan area, comprising 44,444 consecutive patients urgently hospitalized in intensive/cardiovascular care units with a primary diagnosis of ADHF between 2013 and 2021. Clinical phenotypes, i.e., pulmonary congestion or tissue hypoperfusion, were defined through a comprehensive assessment of clinical signs and symptoms, vital signs, and laboratory findings. We assessed the frequency and temporal trends in phenotype-based drug utilization of vasoactive agents and investigated institutional characteristics associated with adopting the phenotype-based approach.

**Results:** Throughout the study period, both overall and phenotype-based vasodilator utilization showed significant declines, with overall usage dropping from 61.4% in 2013 to 48.6% in 2021 (*p for trend* < 0.001). Conversely, no temporal changes were observed in overall inotrope/vasopressor utilization from 24.6% in 2013 to 25.8% in 2021 or the proportion of phenotype-based utilization. Notably, there was considerable variability in phenotype-based drug utilization among hospitals, ranging from 0% to 100%. Particularly, hospitals with a large number of board-certified cardiologists demonstrated reduced phenotype-based vasodilator utilization and phenotypically inappropriate inotrope/vasopressor utilization over time.

**Conclusions:** Substantial variability existed among hospitals in phenotype-based drug utilization of vasoactive agents for ADHF patients, highlighting the need for standardization in their adoption during hospitalization.

**Key Learning Points:** *a. What is known:*

- The absence of standardized drug utilization practices for acute decompensated heart failure contributes to notable variations in treatment approaches across different healthcare facilities, and these facility-specific differences have potential to influence patient outcomes.

*b. What the study adds:*

- This study demonstrated that, using a government-funded multicenter registry of hospitalized patients for acute decompensated heart failure, overall and phenotype-based inotrope/vasopressor utilization was relatively stable between 2013 and 2021, while significant declines in both overall and phenotype-based vasodilator utilization was observed over the same period.
- Significant variability was noted in the extent of phenotype-based drug utilization among hospitals, with the utilization rates ranging from 0% to 100%: Specifically, hospitals with a large number of board-certified cardiologists demonstrated gradual declines in phenotype-based vasodilator utilization and phenotypically inappropriate utilization of inotropes/vasopressors.

## INTRODUCTION

Heart failure (HF) is a global healthcare concern, affecting approximately 64 million individuals worldwide.^1^ Recent data from the National Inpatient Sample in the United States and European surveys indicate a rising trend in HF hospitalizations over the past decade.^2,3^ Similarly, the Japanese Circulations Society reported a significant increase in hospitalized HF patients, surpassing 10,000 new cases annually.^4^ Of note, the economic burden attributed to HF hospitalizations in Japan exceeds that of any other acute cardiovascular disease.^5^

In addition to the increasing incidence and costs, HF remains a significant healthcare challenge characterized by high morbidity and mortality rates. This challenge is particularly pronounced during and following acute exacerbations.^6-8^ While therapeutic options in the acute phase of HF have seen limited evolution over the past two decades, there has been growing interest in tailoring treatment based on the clinical phenotype, integrating vasoactive agents alongside diuretics to manage congestion and tissue hypoperfusion.^9,10^ However, existing guideline recommendations for such interventions remain modest.^11-13^ At present, a considerable degree of heterogeneity exists in the utilization of vasoactive agents.^14^

Research investigating inter-hospital disparities in the use of vasoactive agents in alignment with clinical status is limited. The absence of standardized drug utilization practices contribute to notable variations in treatment approaches across different healthcare facilities,^15^ and these facility-specific differences have potential to influence patient outcomes.^16,17^ This study aimed 1) to elucidate temporal trends in the utilization of vasoactive agents, 2) to assess institutional differences in the utilization of vasoactive agents tailored to the clinical phenotype, considering congestion and tissue hypoperfusion status, and 3) to examine the potential association between institutional variability in these treatment approaches and hospital characteristics among patients hospitalized for acute decompensated heart failure (ADHF).

## METHODS

### Study cohort

The rationale and design of the Tokyo Cardiovascular Care Unit (CCU) Network Database have been described previously.^18,19^ Briefly, the primary objective of the Tokyo CCU Network, funded by the Tokyo Metropolitan Government, was to assess the utilization of emergency medical services, emergency rooms, general intensive care unit (ICU)/CCU, and other acute inpatient care units in the management of acute cardiovascular conditions. This assessment encompassed not only public hospitals but also private hospitals that accept emergency medical services. Additionally, the registry aimed to evaluate the quality of care and identify areas for improvement in the management of these patients. This information was collected to focus on and effectively manage acute cardiovascular care within ICUs/CCUs and was further implemented officially as epidemiological statistics and reports on the medical care, sanitation, and environment in Tokyo, Japan.

As of May 2024, 76 hospitals that serve a population of 13 million individuals in the metropolitan Tokyo area were included in the database. Individual information was recorded in the database by the registry members of each institution, and the final datasets were collected by the scientific committee under conditions of anonymity, according to the ethical guidelines on epidemiological surveys released by the Japanese Ministry of Health, Labor, and Welfare. The study protocol was approved by the institutional review board of each participating hospital and was conducted in accordance with the Declaration of Helsinki. Informed consent was obtained from all patients and/or their legal representatives.

Between January 1, 2013 and December 31, 2021, 44,444 patients hospitalized with a primary diagnosis of ADHF who required for emergent care in the ICU or CCU were registered in the Tokyo CCU Network Database. Notably, patients presenting with acute coronary syndromes were not included in the present analysis. Cardiologists at each hospital made the clinical diagnosis of ADHF based on both signs and symptoms.^20^ Patients who had a concentration of B-type natriuretic peptide (BNP) < 100 pg/mL or N-terminal proBNP < 300 pg/mL at the time of admission,^21^ those who required a maintenance hemodialysis before the index hospitalization, and those who had missing data of an assessment for clinical phenotypes (i.e., congestion and perfusion status) upon presentation were excluded from the analysis (**Figure 1**). As a result, 37,293 patients were included in the present analysis.

**Figure 1.**
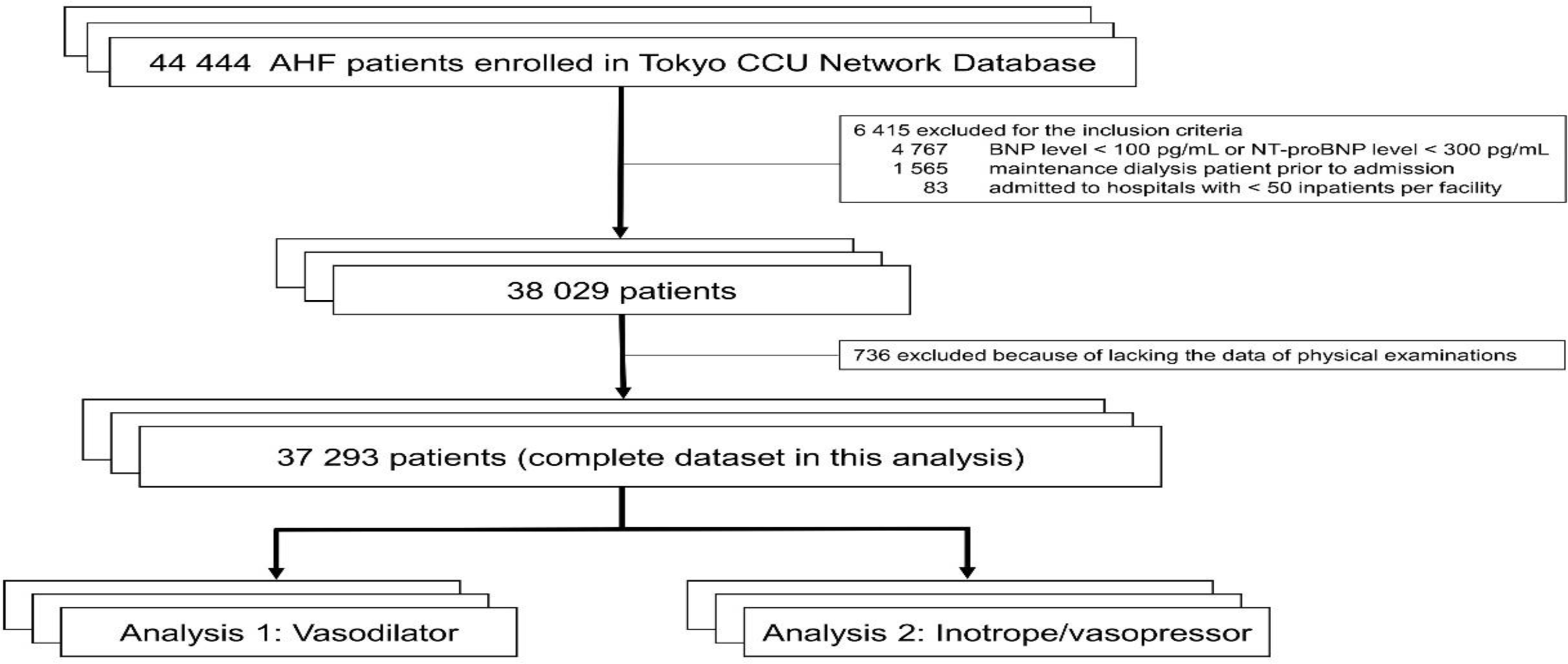
Patient flow chart.

### Definitions of variables and outcomes

The attending cardiologists assessed the presence of clinical signs and symptoms of congestion and tissue hypoperfusion status following the European Society of Cardiology (ESC) guidelines and the Japanese Circulation Society / Japanese Heart Failure Society guidelines during the initial patient assessment.^12,13^ The Nohria and Stevenson classification has been proposed to facilitate a pathophysiology-based intervention for clinicians.^22^ Congestion was accepted by the clinical presence of pulmonary congestion, orthopnea/paroxysmal nocturnal dyspnea, peripheral bilateral edema, jugular venous distention, congested hepatomegaly, gut congestion, ascites and/or hepatojugular reflex. Tissue hypoperfusion was considered to be present if cold sweaty extremities, oliguria, mental confusion, dizziness, and/or narrow pulse pressure (that was clinically ascertained and accepted based on cardiologist perception of weak pulse) were identified.

In this study, we defined clinical phenotypes of pulmonary congestion and tissue hypoperfusion using physical findings, vital signs, and laboratory data: As for pulmonary congestion, 1) physical findings only (i.e., the Nohria and Stevenson classification); 2) physical findings, respiratory rate > 25 breaths per minute, and systolic blood pressure (SBP) > 110 mm Hg based on the 2021 ESC guidelines ^12^; and 3) physical findings, respiratory rate > 30 breaths per minute, and SBP ≥ 160 mm Hg or mean blood pressure (MBP) ≥ 120 mm Hg based on the American College of Cardiology expert consensus.^23^ In addition, we defined tissue hypoperfusion as following: 1) physical findings only; 2) physical findings and vital signs (i.e., proportional pulse pressure < 0.25 or SBP < 90 mm Hg or MBP < 65 mm Hg) ^24^; and 3) physical findings and organ failure (i.e., renal impairment [i.e., creatinine clearance < 60 ml/min] or disturbance of consciousness).^22^ As the Tokyo CCU network database included data on serum lactate levels at initial presentation from 2017 to 2021, we defined tissue hypoperfusion status additionally based on vital signs (i.e., SBP < 90 mm Hg or MBP < 65 mm Hg) and a serum lactate level ≥ 2.0 mmol/L, aligning with the Society for Cardiovascular Angiography and Interventions (SCAI) stage C to E.^25^

Based on pathophysiological conditions and guideline recommendations, the clinically relevant use of intravenous administration of vasoactive agents was defined as follows:

- Clinically appropriate use of vasodilators (i.e., nitrates, carperitide, and nicorandil) was defined to relieve pulmonary congestion. These agents should not be used for patients with cardiogenic shock and/or serious hypotension with SBP of < 90 mm Hg.
- Inotropes and vasopressors (i.e., catecholamines and phosphodiesterase 3 inhibitors) were used to improve peripheral circulatory failure (any signs and symptoms of tissue hypoperfusion). Conversely, the use of inotropes and vasopressors in patients lacking evidence of tissue hypoperfusion was considered to be inappropriate due to the potential for adverse effects such as arrhythmia and myocardial ischemia.

### Statistical analysis

Initially, we analysed the prevalence of patients presenting with pulmonary congestion and tissue hypoperfusion status based on their respective definitions. Subsequently, we provided a descriptive overview of hospitals within the Tokyo Metropolitan area, with a particular focus on the number of board-certified cardiologists per hospital. Hospitals were categorized according to the number of board-certified cardiologists, and patient-level differences in baseline characteristics were assessed using p value and standardized mean difference, given the large sample size. Continuous variables were presented as median and interquartile range (IQR), while categorical variables were presented as frequency and percentage.

Next, we examined the variation in vasoactive agent utilization, specifically phenotype-based utilization of vasodilators and inotropes/vasopressors, across all hospitals according to different definitions of pathophysiological conditions. We described the temporal trends and the utilization of vasoactive agents during the study periods stratified by the presence of pulmonary congestion and tissue hypoperfusion. In addition, we delineated the frequency distribution of phenotype-based drug utilization for vasoactive agents across hospitals as a proportion of their eligible patients.

For the analysis of tissue hypoperfusion using the SCAI classification, missing values of serum lactate levels were addressed using Multivariate Imputation by Chained Equations (MICE). Specifically, we generated five imputed datasets, each through five iterations to ensure robustness in the imputation process.

All probability values were 2-tailed, and values of p < 0.05, were considered statistically significant. All analyses were conducted using the tidyverse (version 2.0.0) and mice (version 3.16.0) in R version 4.2.3 (R Foundation for Statistical Computing, Vienna, Austria, 2008).

## RESULTS

### Clinical phenotypes and various definitions

Of the 37,293 patients included in the study cohort, 33,030 (88.6%) and 7,892 (21.2%) had clinical symptoms and signs of congestion and tissue hypoperfusion status, respectively. The distribution of patients in each definition is shown as **Figure 2** in the Euler diagrams. Among 21,864 patients enrolled between 2017 and 2021, 907 patients (4.1%) were classified as having tissue hypoperfusion status using the SCAI stage C to E.

**Figure 2.**
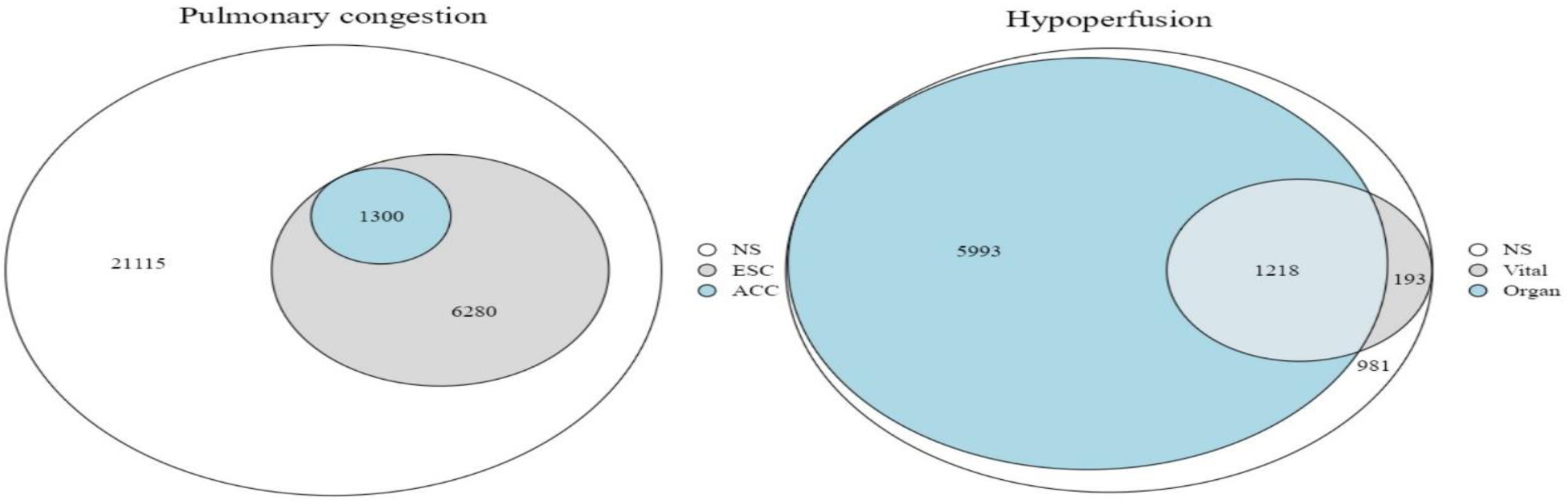
The proportion of each definition of congestion and tissue hypoperfusion. Patients with pulmonary congestion only (excluding a complication of tissue hypoperfusion) was included in the Euler diagram.

### Temporal trends in vasoactive agent usage

While the overall utilization of inotropes/vasopressors relatively stable between 2013 and 2021 (from 24.6% in 2013 to 25.8% in 2021), there was a significant decline in that of vasodilators from 61.4% in 2013 to 48.6% in 2021 (*p for trend* < 0.001) (**Figure S1**). Specifically, there was a marked decrease in the utilization of carperitide from 48.3% in 2013 to 18.5% in 2021, contrasted by a gradual increase in that of nitrates from 27.3% in 2013 to 35.9% in 2021 (both *p for trend* < 0.001) (**Figure 3**). As for inotropes/vasopressors, there was a declining trend in the utilization of dopamine from 7.2% in 2013 to 3.2% in 2021, while that of dobutamine experienced a gradual rise from 15.9% in 2013 to 19.2% in 2021. There were no notable changes in the utilization rates of noradrenaline, adrenaline, and phosphodiesterase inhibitors throughout the study period.

**Figure 3.**
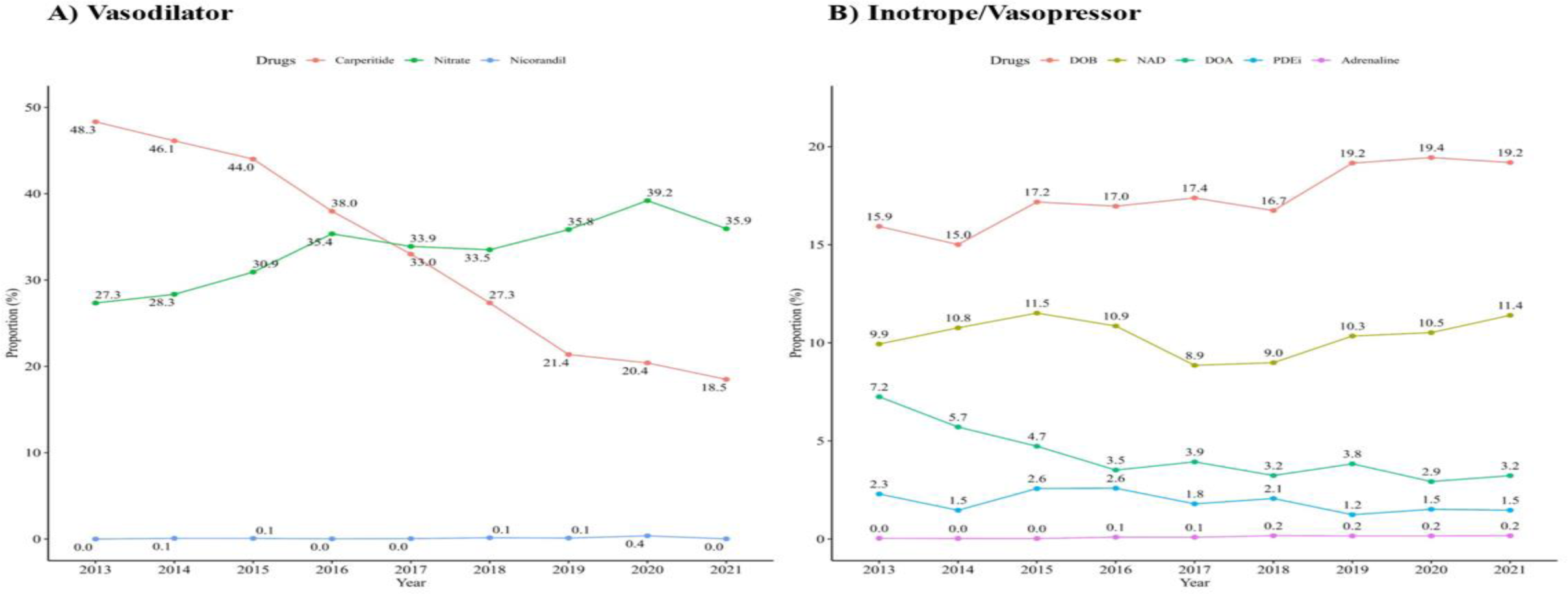
Temporal trends in the utilization of individual vasoactive agents between 2013 and 2021.

No significant temporal changes were observed in the prevalence of inotrope/vasopressor usage according to tissue hypoperfusion status (i.e., phenotype-based drug utilization) over the course of the study period (**Figure S2 and S3**). On the other hand, there was a significant decrease over time in phenotype-based vasodilator utilization according to pulmonary congestion status, even in any definitions (all *p for trend* < 0.001).

### Per hospital use of vasoactive agents

The extent of phenotype-based vasodilator utilization per hospital varied widely, ranging from 0% to 100%, with a median of 57.2% (IQR: 49.3–63.8), 64.7% (IQR: 54.7–74.1), and 72.7% (IQR: 59.2–86.7) by each definition, respectively (**Figure 4A**). Similarly, phenotype-based inotrope/vasopressor utilization varied significantly across hospitals, ranging from 0% to 100%, with a median of 43.9% (IQR: 34.5–55.6), 64.7% (IQR: 51.2–77.8), and 44.0% (IQR: 35.2–57.5) in each definition, respectively (**Figure 4B**).

**Figure 4.**
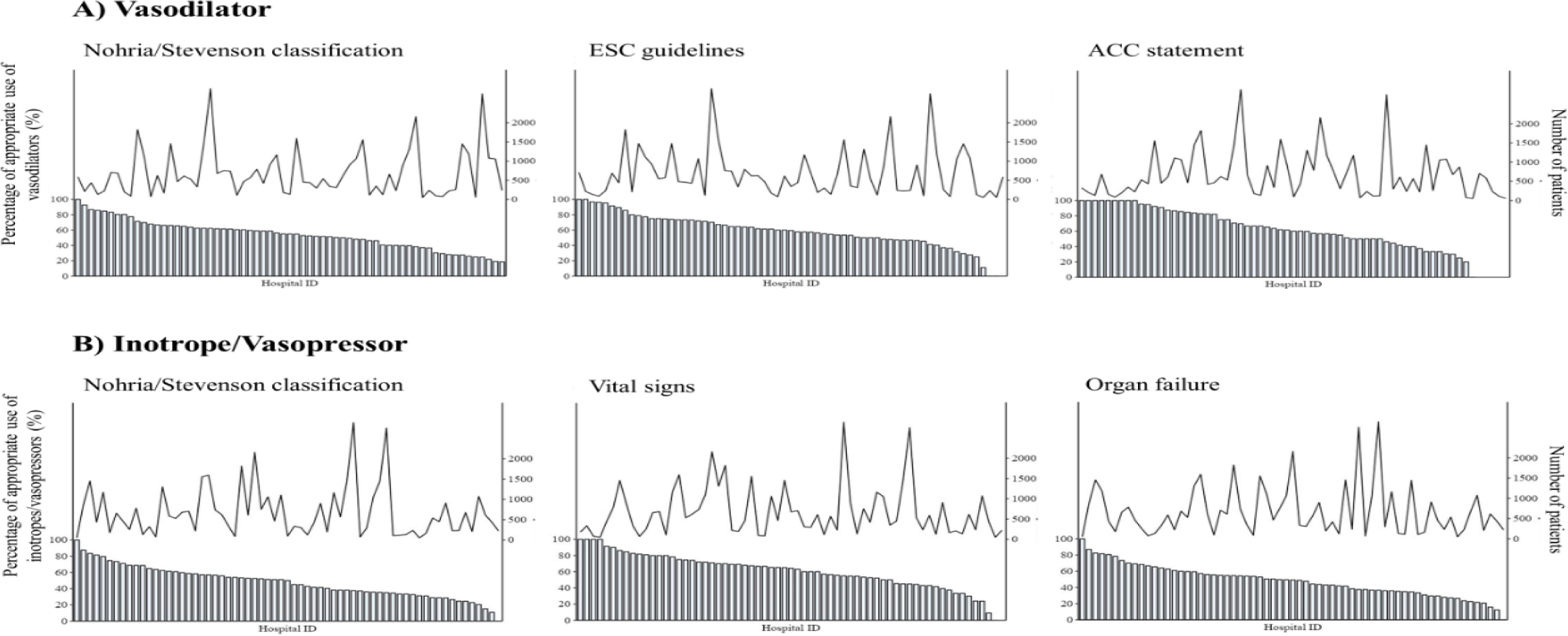
Vasoactive agent utilization, A) vasodilators and B) inotropes/vasopressors, aligned with clinical phenotypes (percentage) ranked-ordered by hospitals and the number of patients by hospitals (number). The bar graph shows the percentage of pathophysiology-matched drug uses at each hospital, and the line graph shows the number of patients at each hospital.

### Patient characteristics

**Table 1** shows the patient characteristics categorized by hospital groups according to the mean number of board-certified cardiologists (n < 5 vs. n ≥ 5). Notably, there were no significant inter-hospital differences in the prevalence of pulmonary congestion and tissue hypoperfusion status, with exception of physical findings related to pulmonary congestion. Furthermore, there were no discrepancies observed in other patient characteristics, aside from the utilization of vasodilators. Additionally, there was no difference in the in-hospital mortality rates between the two groups. It is worth mentioning that hospitals with a large number (n ≥ 5) of board-certified cardiologists tended to handle a greater number of HF cases per hospital compared to those with fewer (n < 5) board-certified cardiologists, as illustrated in **Figure S4**.

**Table 1.** Patient backgrounds according to the number of board-certified cardiologists. * 36,637 patients were analyzed after excluding 656 patients enrolled in five hospitals that did not have data on the number of board-certified cardiologists. Abbreviations: SMD: standard mean difference, SBP: systolic blood pressure, COPD: chronic obstructive pulmonary disease, HF, heart failure, MI: myocardial infarction, PCI: percutaneous coronary intervention, CABG: coronary artery bypass grafting, NT-proBNP: N-terminal pro-B-type natriuretic peptide, ACEi: angiotensin-converting enzyme inhibitors, ARB: angiotensin receptor antagonist, IV: intravenous, IABP: intra-aortic balloon pump, VA-ECMO: veno-arterial extracorporeal membrane oxygenation, ICU: intensive care unit, CCU: cardiovascular care unit.

| Value | Hospitals with a small number of<br>board-certified cardiologists<br>(n = 17,001) | Hospitals with a large number of<br>board-certified cardiologists<br>(n = 19,636) | P value | SMD |
| --- | --- | --- | --- | --- |
| Age, years | 80 (70–87) | 80 (71–87) | 0.196 | 0.009 |
| Female, n (%) | 7431 (43.7) | 8573 (43.7) | 0.972 | <0.001 |
| Body mass index, kg/m<sup>2</sup> | 21.9 (19.8–24.8) | 21.9 (19.3–24.6) | <0.001 | 0.054 |
| SBP, mm Hg | 143 (120–168) | 142 (120–168) | 0.087 | 0.007 |
| Heart rate, bpm | 101 (83–120) | 100 (81–120) | 0.026 | 0.022 |
| Respiratory rate, rpm | 24 (19–26) | 24 (19–28) | 0.014 | 0.028 |
| Oxygen saturation, % | 93 (86–97) | 93 (85–97) | 0.146 | 0.033 |
| Left ventricular ejection fraction, % | 40 (30–50) | 40 (30–51) | 0.004 | 0.038 |
| Pulmonary congestion |  |  |  |  |
| Physical findings, n (%) | 13922 (81.8) | 14699 (74.9) | <0.001 | 0.170 |
| ESC guidelines, n (%) | 3468 (20.4) | 3904 (19.9) | 0.234 | 0.013 |
| ACC statements, n (%) | 669 (3.9) | 804 (4.1) | 0.447 | 0.008 |
| Tissue hypoperfusion |  |  |  |  |
| Physical findings, n (%) | 3895 (22.9) | 4647 (23.7) | 0.085 | 0.018 |
| Vital signs, n (%) | 822 (4.8) | 949 (4.8) | 0.999 | <0.001 |
| Organ failure, n (%) | 3283 (19.3) | 3975 (20.2) | 0.025 | 0.024 |
| Hypertension, n (%) | 11123 (65.4) | 12480 (63.6) | <0.001 | 0.038 |
| Diabetes mellitus, n (%) | 5992 (35.2) | 6512 (33.2) | <0.001 | 0.043 |
| Stroke, n (%) | 1972 (11.6) | 2523 (12.8) | <0.001 | 0.038 |
| Peripheral artery disease, n (%) | 609 (3.6) | 648 (3.3) | 0.15 | 0.015 |
| COPD, n (%) | 1072 (6.3) | 1154 (5.9) | 0.094 | 0.018 |
| Atrial fibrillation, n (%) | 6736 (39.6) | 7653 (39.0) | 0.227 | 0.013 |
| Previous hospitalization for HF, n (%) | 6718 (39.5) | 7706 (39.2) | 0.636 | 0.005 |
| Previous MI, n (%) | 2877 (16.9) | 3292 (16.8) | 0.717 | 0.004 |
| Previous PCI, n (%) | 2774 (16.3) | 3208 (16.3) | 0.949 | 0.001 |
| Previous CABG, n (%) | 1180 (6.9) | 1151 (5.9) | <0.001 | 0.044 |
| Etiology of HF |  |  | <0.001 | 0.098 |
| Coronary artery disease, n (%) | 4294 (25.2) | 4300 (21.9) |  |  |
| Dilated cardiomyopathy, n (%) | 1611 (9.5) | 2165 (11.0) |  |  |
| Valvular disease, n (%) | 3344 (19.7) | 3645 (18.6) |  |  |
| Others, n (%) | 7762 (45.6) | 9526 (48.5) |  |  |
| Hemoglobin, g/dL | 11.8 (10.2–13.5) | 11.9 (10.2–13.7) | <0.001 | 0.04 |
| Blood urea nitrogen, mg/L | 24.3 (17.9–36.0) | 24.5 (18.0–36.7) | 0.182 | 0.018 |
| Creatinine, mg/mL | 1.1 (0.8–1.6) | 1.1 (0.9–1.7) | <0.001 | 0.003 |
| Albumin, mg/L | 3.5 (3.1–3.8) | 3.5 (3.1–3.8) | 0.474 | 0.007 |
| NT-proBNP, pg/mL | 3421 (1650–7800) | 2979 (1451–6637) | <0.001 | 0.004 |
| Diuretic at admission, n (%) | 7847 (46.1) | 9249 (47.1) | 0.064 | 0.02 |
| ACEi/ARB at admission, n (%) | 6581 (38.7) | 7056 (35.9) | <0.001 | 0.057 |
| Beta blocker at admission, n (%) | 6509 (38.3) | 7715 (39.3) | 0.045 | 0.021 |
| IV vasodilator use, n (%) | 10306 (60.6) | 10558 (53.8) | <0.001 | 0.138 |
| IV inotrope/vasopressor use, n (%) | 4112 (24.2) | 4980 (25.4) | 0.009 | 0.028 |
| IABP use, n (%) | 474 (2.8) | 420 (2.1) | <0.001 | 0.042 |
| Impella use, n (%) | 37 (0.2) | 25 (0.1) | 0.049 | 0.022 |
| VA-ECMO use, n (%) | 122 (0.7) | 116 (0.6) | 0.151 | 0.016 |
| Patient number in public hospitals, n (%) | 2727 (17.2) | 7592 (38.7) | <0.001 | 0.494 |
| Patient number in tertiary hospitals, n (%) | 5855 (36.9) | 12527 (63.8) | <0.001 | 0.559 |
| ICU/CCU bed, n | 6 (5–10) | 6 (4–8) | <0.001 | 0.254 |
| Board-certified cardiologist, n | 3 (3–4) | 7 (6–8) | <0.001 | 2.294 |
| In-hospital mortality, n (%) | 1527 (9.0) | 1562 (8.0) | <0.001 | 0.037 |

### Hospital characteristics and treatment approaches

We assessed the relationships between phenotype-based drug utilization and various hospital characteristics, including hospital types (i.e., tertiary care provider, public/private hospitals), as well as the number of ICU/CCU beds and board-certified cardiologists. While no clear relationships emerged between phenotypically appropriate utilization of inotropes/vasopressors and hospital characteristics, hospitals with a large number of board-certified cardiologists demonstrated a significant association with reduced frequencies of phenotype-based vasodilator utilization (*p for trend* < 0.001) (**Figure S5**). Furthermore, a downward trend in the phenotypically inappropriate utilization of inotropes/vasopressors for tissue hypoperfusion status was observed in hospitals with a large number of board-certified cardiologists (*p for trend* < 0.001) (**Figure S6**). On a sensitivity analysis using the SCAI classification, there were no significant temporal changes in both phenotypically appropriate and inappropriate utilization of inotropes/vasopressors.

## DISCUSSION

Using a government-funded acute cardiovascular care database in Tokyo, Japan, we examined the utilization patterns of vasodilators and inotropes/vasopressors among patients admitted to ICUs/CCUs with ADHF. Notably, vasodilators were frequently administered, particularly among approximately half of the cases presenting with pulmonary congestion, while inotropes/vasopressors were employed in a limited subset of patients presenting with tissue hypoperfusion. Our analysis revealed that the utilization of inotropes/vasopressors remained relatively unchanged during the study period, while there was a significant decline in both overall and phenotype-based utilization of vasodilators over the same time frame. However, substantial variability existed among practice patterns with regard to the use of vasoactive agents across hospitals. Specifically, we observed gradual declines in phenotype-based vasodilator utilization and phenotypically inappropriate utilization of inotropes/vasopressors among hospitals with many a large number of board-certified cardiologists.

In our study, established clinical criteria were employed to define pulmonary congestion and tissue hypoperfusion status, yet the observed trends remained consistent irrespective of the specific definition applied. Notably, clinicians did not base their decisions regarding drug usage solely on physical findings, as indicated by the Nohria-Stevenson classification. Instead, treatment decisions appeared to be predominantly influenced by a combination of vital signs and laboratory findings, as evidenced by the higher proportion of phenotype-based treatment observed with more detailed definitions. Clinicians may be cautious in prescribing vasodilators, recognizing their potential to induce excessive hypotension, particularly in patients presenting with normotension or signs suggestive of tissue hypoperfusion. ^26-28^ Surprisingly, inotropes/vasopressors were not administered in nearly half of the cases, even in the presence of tissue hypoperfusion status at presentation, with other treatment modalities such as diuretics and vasodilators being preferred. This observation may reflect the limitations associated with relying solely on physical and vital signs for assessing tissue hypoperfusion status. Of note, prior research has highlighted the lack of reliability in using physical findings alone to predict tissue hypoperfusion status.^29^ Additionally, the lower variability observed in the use of inotropes/vasopressors compared to vasodilators across each hospital may be attributed to a shared perspective derived from previous studies, which emphasize the importance of minimizing the use of inotropes whenever possible.^12,30^

The use of vasoactive agents in the management for ADHF varies substantially by the region. In a multinational observational study,^14^ vasodilator usage for ADHF patients was reported to be infrequent, aligning with the limited endorsement of these drugs in European and American clinical practice guidelines.^11,12^ Conversely, the West-Pacific region showed a higher prevalence of vasodilator usage,^14^ irrespective of the recommendations within the clinical practice guidelines.^30^ A joint analysis of three large-scale cohort studies in Japan revealed that intravenous vasodilators were frequently used in over 50% of hospitalized HF patients between 2007 and 2015,^8^ partially attributed to the strong recommendation for intravenous vasodilators to relieve signs and symptoms of pulmonary congestion (Class IIa recommendation) as per domestic clinical practice guidelines.^13^ However, our analysis indicated a consistent decline in the utilization of vasodilators, particularly carperitide, in recent years. This decline may be influenced by numerous reports from Japan, including our own, demonstrating unfavorable associations between carperitide usage and clinical outcomes.^26,31,32^ Furthermore, a revision of the Japanese HF guidelines took place in 2017. These updated guidelines emphasized pathophysiological phenotype-based therapies to a greater extent than the previous 2011 guidelines.^13^ This shift in emphasis may have had an impact on the frequency of vasodilator usage, particularly resulting in decreased utilization of carperitide, aimed at achieving diuretic effects.

We found the gradual decrease in the utilization of dopamine, contrasted by the increase in the utilization of dobutamine. This may be attributed to the impact of negative findings from previous trials and guidance provided by domestic clinical practice guidelines.^13,33,34^ In our study, approximately 25% of cases received inotropes/vasopressors, which represents a higher frequency compared to previous Japanese studies.^8,35,36^ One potential explanation for this difference is the inclusion of critically ill patients admitted to the ICU/CCU in our registry, which likely contributed to the higher observed mortality rate compared to other studies ^8,35,36^

Importantly, our study revealed substantial disparities and variances in vasoactive agent utilization across different hospitals, suggesting potential reflection of individual hospital practices. These discrepancies may be influenced by the lack of uniformity in recommendations and treatment strategies found in clinical practice guidelines for HF, both in Japan and internationally.^11-13^ Furthermore, differences in hospital characteristics, particularly the number of board-certified cardiologists, could contribute to these variations. Our data indicated that hospitals with a large number of board-certified cardiologists often manage a greater volume of HF patients per hospital, which may influence treatment practices. It is noteworthy that all hospitals within the Tokyo CCU network are capable of providing high-quality acute cardiovascular care, including primary coronary intervention for acute myocardial infarction within one hour of presentation.^18^ Consequently, there were no significant differences in patient backgrounds among hospitals. Following initial treatments, if necessary, patients are transferred to specialized facilities equipped to manage patients with advanced HF.

Our study has several limitations that need to be considered. Firstly, we were unable to gather serial data on physical and laboratory findings throughout the index hospitalization. Additionally, we did not collect data on SBPs following initial treatments, including the intravenous administration of vasodilators, which is crucial for assessing the incidence of hypotension, a factor that can worsen patient prognosis. Secondly, the strength of our study lies in the prospective collection of data using the Nohria and Stevenson classification, although it is essential to acknowledge that the physical examination findings are at the discretion of the attending cardiologists, which introduces the potential for inter-rater variability. Thirdly, we lacked information regarding the sublingual administration of vasodilators, such as nitroglycerin, in the emergency department. It is conceivable that some hypertensive patients who did not receive intravenous vasodilators might have received sublingual treatment. Finally, it is worth noting that pulmonary artery catheterization (PAC) provides a comprehensive hemodynamic assessment, but this database does not included data on whether this procedure is performed or its associated parameters. Our primary focus was to examine the association between initial drug treatment and clinical phenotypes at presentation. Including PAC data may present its own challenges, as PAC is typically considered for patients who do not respond to initial treatment. Furthermore, the use of PAC itself implies pathophysiologically phenotype-based treatment, which naturally entails higher rates of phenotype-based drug utilization. These could introduce a bias into our study, potentially leading to the result that hospitals with higher PAC use have higher rates of phenotype-based treatment.

Our study demonstrated that, despite existing recommendations that advocate for the selection of drugs based on underlying pathophysiological factors, only half of the patients received treatments aligned with pathophysiologically specific conditions. Furthermore, our investigation has revealed substantial variability in drug utilization patterns across different healthcare facilities. Our findings emphasize the necessity for further research and assessment to gain a deeper understanding of the influence of tailored drug therapies on patient outcomes. Additionally, it underscores the need to identify strategies for healthcare facilities to standardize and optimize their treatment approaches for HF, with the ultimate goal of improving patient care and outcomes in these critical settings.

## Acknowledgement

The authors would like to thank Ms. Kozue Murayama and Ms. Nobuko Yoshida and the members of Tokyo CCU network scientific committee for their important contributions.

## Data availability

The anonymized data that support the findings of this study are available from the corresponding author for reasonable requests.

